# Effects of COVID-19 mRNA vaccination on HIV viremia and reservoir size

**DOI:** 10.1101/2023.10.08.23296718

**Authors:** Maggie C. Duncan, F. Harrison Omondi, Natalie N. Kinloch, Hope R. Lapointe, Sarah Speckmaier, Nadia Moran-Garcia, Tanya Lawson, Mari L. DeMarco, Janet Simons, Daniel T. Holmes, Christopher F. Lowe, Nic Bacani, Paul Sereda, Rolando Barrios, Marianne Harris, Marc G. Romney, Julio S.G. Montaner, Chanson J. Brumme, Mark A. Brockman, Zabrina L. Brumme

## Abstract

**Objective:** The immunogenic nature of COVID-19 mRNA vaccines led to some initial concern that these could stimulate the HIV reservoir. We analyzed changes in plasma HIV loads (pVL) and reservoir size following COVID-19 mRNA vaccination in 62 people with HIV (PWH) receiving antiretroviral therapy (ART), and analyzed province-wide trends in pVL before and after the mass vaccination campaign.

**Design:** Longitudinal observational cohort and province-wide analysis.

**Methods:** 62 participants were sampled pre-vaccination, and one month after their first and second COVID-19 immunizations. Vaccine-induced anti-SARS-CoV-2-Spike antibodies in serum were measured using the Roche Elecsys Anti-S assay. HIV reservoirs were quantified using the Intact Proviral DNA Assay; pVL were measured using the cobas 6800 (LLOQ:20 copies/mL). The province-wide analysis included all 290,401 pVL performed in British Columbia, Canada between 2012-2022.

**Results:** Pre-vaccination, the median intact reservoir size was 77 (IQR:20-204) HIV copies/million CD4+ T-cells, compared to 74 (IQR:27-212) and 65 (IQR:22-174) post-first and -second dose, respectively (all comparisons p>0.07). Pre-vaccination, 82% of participants had pVL<20 copies/mL (max:110 copies/mL), compared to 79% post-first dose (max:183 copies/mL) and 85% post-second dose (max:79 copies/mL) (p>0.4). The magnitude of the vaccine-elicited anti-SARS-CoV-2-Spike antibody response did not correlate with changes in reservoir size nor detectable pVL frequency (p>0.6). We found no evidence linking the COVID-19 mass vaccination campaign to population-level increases in detectable pVL frequency among all PWH in the province, nor among those who maintained pVL suppression on ART.

**Conclusion:** We found no evidence that COVID-19 mRNA vaccines induced changes in HIV reservoir size nor plasma viremia.

## Introduction

The mass rollout of safe and effective SARS-CoV-2 mRNA vaccines was critical in combatting the COVID-19 pandemic. As people with HIV (PWH) are at a higher risk of severe COVID-19 outcomes [1–3], it was particularly important for this group to be vaccinated, and a large body of evidence now reassuringly confirms that PWH receiving suppressive antiretroviral therapy (ART) generally mount robust immune responses to COVID-19 vaccination [4–12]. Initially however, COVID-19 vaccine confidence was typically lower among PWH compared to the general population [13,14], with possible effects of vaccination on viral rebound cited among the concerns [15,16]. Such concerns are not entirely unfounded, as some vaccines, including those against influenza, Hepatitis B, and pneumococcus can induce HIV transcription, leading to transient increases in plasma HIV RNA levels [17–21].

The immunogenic nature of mRNA vaccines, which elicit strong humoral and cell-mediated immune responses by harnessing innate detectors of single-stranded viral RNA [22,23], led to some initial concerns that these might induce HIV expression, and possibly viral release, from the reservoir [24]. This could theoretically occur via direct stimulation of reservoir cells that recognize the vaccine antigen, or through a generalized inflammatory response with cytokine production that could transiently promote HIV gene expression. Indeed, reports of increased HIV viremia in individuals receiving ART following COVID-19 mRNA vaccination have emerged [25–27], though other studies have observed no such effects [28–30].

Existing studies however have been relatively modestly sized. A recent analysis of 35 PWH, which included 15 participants from the present cohort, reported that the frequency of Nef-specific CD8+ T cells transiently increased after the initial COVID-19 mRNA vaccine dose, consistent with immune sensing of reactivated reservoir cells, but plasma viremia was not investigated and no significant changes in reservoir size were observed in the subset of 13 participants analyzed for this outcome [31]. Another analysis of 25 PWH reported no significant changes in reservoir size post-COVID-19 vaccination [29]. A recent analysis of 68 PWH reported a gradual yet not statistically significant increase in pVL after the second vaccine dose with no obvious effects on reservoir size, but nearly half of participants received the viral vectored ChAdOx1 vaccine (which may be less likely to modulate the reservoir), and pVL and reservoir data were available for fewer than two-thirds of the cohort [27]. To our knowledge, no studies have investigated the effects of COVID-19 mRNA vaccination on the reservoir in an observational cohort while also analyzing population-level trends in pVL test results in a large geographic region following mass COVID-19 vaccination.

Here, we analyzed changes in pVL and reservoir size following the first and second COVID-19 mRNA vaccine doses in 62 PWH receiving ART. Using a longitudinal dataset that captured all PWH in British Columbia (BC), Canada, we also investigated whether the frequency of detectable HIV RNA test results increased at the population level following the mass administration of first, second and booster COVID-19 vaccine doses in the province.

## Methods

### Cohort and specimen collection

Our cohort of PWH on ART, established at the outset of the mass COVID-19 vaccination campaign in BC, has been described previously [6]. The present analysis includes all PWH who provided a pre-vaccination sample and who received two mRNA vaccine doses (either BNT162b2 or mRNA-1273). Serum, plasma and peripheral blood mononuclear cells (PBMC; isolated by density gradient separation and cryopreserved at −150°C until analysis) were collected pre-vaccination, and again one month after the first and second vaccine doses.

### Ethics Approval

The cohort study was approved by the University of British Columbia/Providence Health Care and Simon Fraser University Research Ethics Boards. All participants provided written informed consent. The BC Centre for Excellence in HIV/AIDS (BC-CfE) Drug Treatment Program (DTP), the source of the province-wide pVL dataset, is a provincially-funded clinical registry mandated to: i) deliver health care to individuals living with HIV and related diseases, or at risk of HIV infection, ii) implement and support public health initiatives to curb HIV/AIDS, iii) monitor and evaluate these health care programs, and iv) support related knowledge translation and educational programs. As a result, the requirement for REB review of the province-wide pVL analysis was waived by the Providence Health Care/University of British Columbia REB.

### Anti-SARS-CoV-2 antibody assays

Total binding antibodies against SARS-CoV-2 nucleocapsid (N) and spike receptor binding domain (RBD) in serum were measured using the Roche Elecsys Anti-SARS-CoV-2 and Anti-SARS-CoV-2 S assays, respectively. Both are electro-chemiluminescence sandwich immunoassays. The presence of anti-N antibodies identified participants with prior SARS-CoV-2 infection. The S assay reports results in arbitrary units/mL (U/mL) calibrated against an external standard, where the measurement range is from 0.4-25,000 U/mL.

### Plasma HIV RNA quantification

Plasma HIV RNA levels were quantified using the cobas Ampliprep/Taqman HIV-1 Test v2.0 (during the period March 7, 2012 – June 4, 2018) or the cobas HIV-1 Test on a cobas 6800 (from June 5, 2018 – present; Roche Diagnostics). The lower limit of quantification (LLOQ) of this test is 20 HIV RNA copies/mL. This threshold defined undetectable viremia, unless otherwise indicated.

### HIV Reservoir Quantification

CD4+ T-cells were isolated from PBMCs via negative selection using the EasySep Human CD4+ T-cell Enrichment Kit (STEMCELL Technologies). Median CD4+ T-cell purity, assessed flow cytometrically post-isolation, was 97%. Genomic DNA was extracted from a median of 2.9 (Interquartile Range [IQR] 2.2-3.8) million CD4 + T-cells using the QIAamp DNA Mini Kit (QIAGEN). HIV reservoir quantification was performed using the Intact Proviral DNA Assay (IPDA) [32] as described previously [33]. Briefly, this droplet digital PCR assay distinguishes genomically-intact proviruses from the vast background of defective ones by simultaneously targeting two HIV regions, the Packaging Signal (Ψ) near the 5’ end of the viral genome and the Rev Responsive Element (RRE) within Envelope (*env*), that together strongly predict genomic intactness. An unlabeled competitive RRE probe specific for hypermutated proviruses ensures that these are not counted as intact.

Occasionally, the published IPDA primers/probes fail to detect a participant’s proviral pool due to sequence polymorphism [33], which occurred in 14 (23%) of participants. For these, we employed a secondary *env* reaction [33] or custom autologous primers/probes. The assay also quantifies human genomic DNA in an independent parallel reaction, using primer/probe sets in the human RPP30 gene that are spaced the same distance apart as the HIV target regions. This spacing also allows each sample’s data to be corrected for the DNA shearing that occurs during extraction (by measuring the frequency whereby the RPP30 targets are decoupled). The assay reports the number of intact HIV genomes (those positive for both Ψ and RRE), as well as the overall number of proviral DNA copies (those positive for at least one target), per million CD4+ T-cells.

A median of 290,000 (IQR 255,000-325,000) cells were assayed per participant across four replicate wells, which were merged to generate the final reservoir measurement. Genomic DNA from J-Lat 9.2 cells, which harbor a single integrated copy of replication-incompetent HIV per cell, was used as a positive control, while genomic DNA from donors without HIV, and water, were used as negative controls. Droplets were read using the QX200 Droplet Reader (BioRad) and analysed using QuantaSoft (BioRad, version 1.7.4). Wells containing fewer than 10,000 droplets were excluded from analysis. The median DNA shearing was 0.38 (IQR 0.35-0.39), well within the acceptable range [32].

### Temporal analysis of HIV plasma viral load test results in British Columbia

The BC Centre for Excellence in HIV/AIDS (BC-CfE) provides care and treatment for all PWH in the province. The BC-CfE’s Drug Treatment Program (DTP) database captures all HIV plasma viral load (pVL) tests and ART information for all PWH in BC. We analyzed all 290,401 pVL tests performed between January 2012 and December 2022 to investigate whether the frequency of detectable pVL test results increased following each “wave” of mass COVID-19 vaccination in the province. Though the LLOQ of the pVL assay is 20 copies/mL, the results are clinically reported (and thus stored in the DTP database) with a LLOQ of 40 copies/mL. COVID-19 vaccine distribution data for BC up to December 2022 (where 97% of vaccines administered were mRNA) [34] were retrieved from the Public Health Agency of Canada and COVID-19 Vaccine Tracker [34–36].

### Statistical Analysis

Comparisons of continuous variables between groups were performed using the Mann-Whitney U-test (for unpaired data) or Wilcoxon test (for paired data). Correlations between continuous variables were performed using Spearman’s correlation. Frequency comparisons were performed using the χ^2^ test. Where appropriate, multiple comparisons were addressed using a false-discovery rate (q-value) approach [37]. All statistical tests were performed in using R (version 4.3.1).

## Results

### Participant characteristics

The 62 PWH participants in the observational cohort study were a median 43 years old and 55 (89%) were male. Participants had been receiving ART for a median of 6 years, with 74% on integrase inhibitor-based ART at enrolment (**Table 1**). The most recent CD4+ T-cell count, measured a median of 40 (IQR 15-159) days before enrolment, was 725 (IQR 475-915; range 130-1800) cells/mm^3^. The estimated nadir CD4+ T-cell count, recorded a median of 5.6 (IQR 2.8-13) years before enrolment, was 305 (IQR 160-499; range <9-970) cells/mm^3^. At the baseline (pre-vaccination) visit, 51 (82%) of participants had pVL below the LLOQ of 20 copies HIV RNA/mL (the highest pVL observed at baseline was 110 copies/mL). Overall, 69% of participants received two doses of the BNT162b2 COVID-19 mRNA vaccine, 26% received two doses of mRNA-1273, and 5% received a mixed mRNA regimen. Of note, the interval between first and second COVID-19 doses was extended to up to 112 days in Canada due to initially limited vaccine supplies in the country [38]. The vast majority (57/62; 92%) of participants remained COVID-19 naive throughout follow-up, four experienced COVID-19 before vaccination, and one acquired COVID-19 between the first and second vaccine doses.

**Table 1:**

| Characteristic | N = 62 |
| --- | --- |
| Age at enrolment in years, median (IQR) | 43 (35, 56) |
| Sex at birth |  |
| Male, n (%) | 55 (89%) |
| Female, n (%) | 7 (11%) |
| Nadir CD4+ T-cell count in cells/mm<sup>3</sup>, median (IQR) | 305 (160, 498) |
| Baseline<sup>a</sup> CD4+ T-cell count in cells/mm<sup>3</sup>, median (IQR) | 725 (475, 915) |
| Baseline<sup>a</sup> plasma viral load in copies HIV RNA/mL, median (IQR) | <20 (<20, <20) |
| Years on ART, median (IQR) | 6 (3, 14) |
| Current ART regimen |  |
| INSTI-based, n (%) | 46 (74%) |
| NNRTI-based, n (%) | 6 (9.7%) |
| PI-based, n (%) | 5 (8.1%) |
| Intensive<sup>a</sup>, n (%) | 4 (6.5%) |
| CCR5 antagonist-based, n (%) | 1 (1.6%) |
| Initial COVID-19 vaccine regimen |  |
| BNT162b2 + BNT162b2, n (%) | 43 (69%) |
| mRNA-1273 + mRNA-1273, n (%) | 16 (26%) |
| BNT162b2 + mRNA-1273, n (%) | 3 (4.8%) |
| COVID-19 exposure history |  |
| COVID-19 naive throughout follow-up, n (%) | 57 (92%) |
| COVID-19-experienced prior to vaccination, n (%) | 4 (6.5%) |
| COVID-19 between first and second vaccine doses, n (%) | 1 (1.6%) |

<sup>a</sup> At study entry (*i.e.* prior to COVID-19 vaccination)
<sup>b</sup> Regimen containing at least two of the following drug classes: INSTI, NNRTI, PI, CCR5 antagonist

### No evidence that COVID-19 mRNA vaccination induces detectable viremia

HIV pVL testing was performed at the baseline visit, which occurred a median of 12 (IQR 3-26) days prior to vaccination, one month (a median of 31 [IQR 29-33] days) after the first vaccine dose, and again one month (a median of 30 [IQR 29-30] days) after the second vaccine dose (**Fig. 1**). At baseline, 82% (51/62) of participants had a pVL <20 copies/mL, where the highest observed value was 110 copies/mL. One month after the first vaccine dose, 79% (49/62) of participants had pVL <20 copies/mL (highest value 183 copies/mL), a difference that was not statistically significant from baseline (Wilcoxon paired test; p=0.46). One month after the second dose, three participants had temporarily discontinued ART or missed the visit, leaving 59 participants for analysis. Of these, 85% (50/59) had pVL <20 copies/mL (highest value 79 copies/mL), which was not significantly different compared to baseline (p=0.81), nor one month post-first dose (p=0.88). Using a pVL <50 copies/mL threshold produced consistent results: at baseline, 94% (58/62) of participants had a pVL <50 copies/mL, compared to 92% (57/62) one month after the first vaccine dose, and 93% (55/59) after the second (Chi-squared p=0.93). At no point did any participant experience virologic failure (defined as >200 copies/mL [39,40]). Results also remained consistent after excluding visits from participants who had experienced COVID-19 (all p>0.59; not shown). Stratification of the data by sex, COVID-19 vaccine regimen, and ART regimen similarly produced no statistically significant differences in pVL between baseline and post-vaccination visits for any subgroup (all p>0.08; q>0.78; not shown).

**Figure 1:**
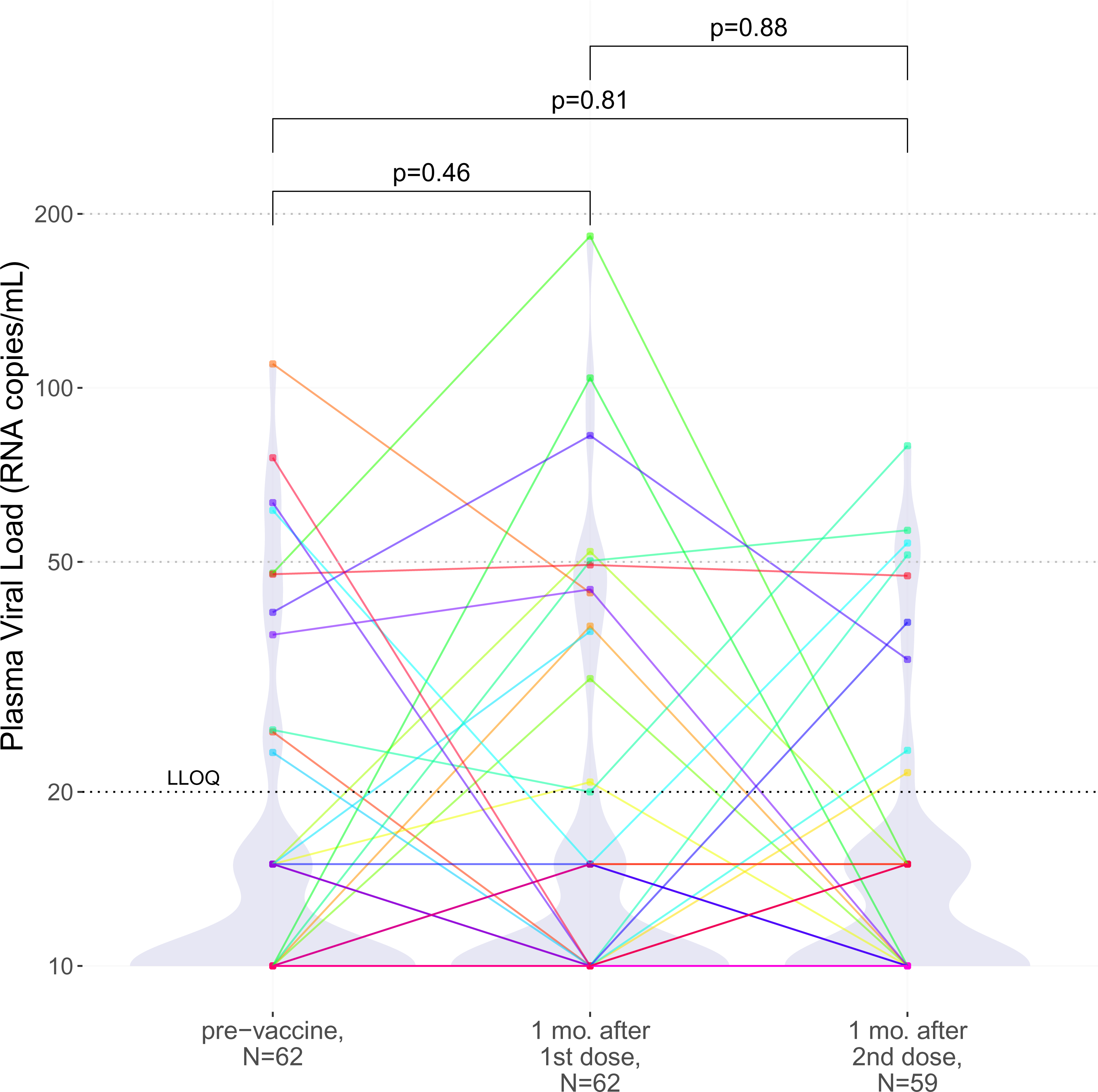
Plasma HIV loads following one- and two-dose COVID-19 vaccination. HIV plasma viral loads prior to vaccination (left), one month after the first dose (middle), and one month following the second dose (right). A black dashed line indicates the assay LLOQ of 20 HIV copies/mL, while light grey dashed lines indicate clinically relevant pVL thresholds of 50 and 200 HIV copies/mL. For graphing purposes, undetectable viral loads were plotted as 10 HIV copies/mL, while viral loads that were detectable yet below the LLOQ were plotted as 15 HIV copies/mL. As the vast majority of pVL measurements were below the LLOQ, violin plots help visualize the data distribution. Each participant is identified by a unique color that is consistent throughout all figures. P-values were calculated using the Wilcoxon sum rank test for paired data.

### No changes in HIV reservoir size after COVID-19 mRNA vaccination

To determine whether COVID-19 mRNA vaccination induced changes in HIV reservoir size (defined as genome-intact proviral load) or total HIV DNA load, we quantified the number of intact, defective and total proviral copies per million CD4+ T-cells (**Fig. 2**) [32]. At baseline, the median number of intact HIV copies per million CD4+ T-cells was 77 (IQR 20-204). One month following the first vaccine dose it was 74 (IQR 27-212), a difference that was not statistically significant (Wilcoxon paired test, p=0.64) (**Fig. 2a**). One month following the second vaccine dose, the median intact reservoir size was 65 (IQR 22-174), which was not significantly different from baseline (p=0.32), nor one month post-first dose (p=0.07) (**Fig. 2a**). Likewise, 5’-defective, 3’-defective, and total proviral burdens did not change significantly between baseline and either post-vaccine visit (**Fig. 2a-2d**; all p>0.08). These results remained consistent after excluding data from participants who experienced COVID-19 (all p>0.07; not shown). Stratification of the data by sex, vaccine, and ART regimen similarly produced no statistically significant differences in intact reservoir size, nor in the total, 5’-defective nor 3’-defective proviral burdens for any subgroup, after adjusting for multiple comparisons (all q>0.24; not shown).

**Figure 2:**
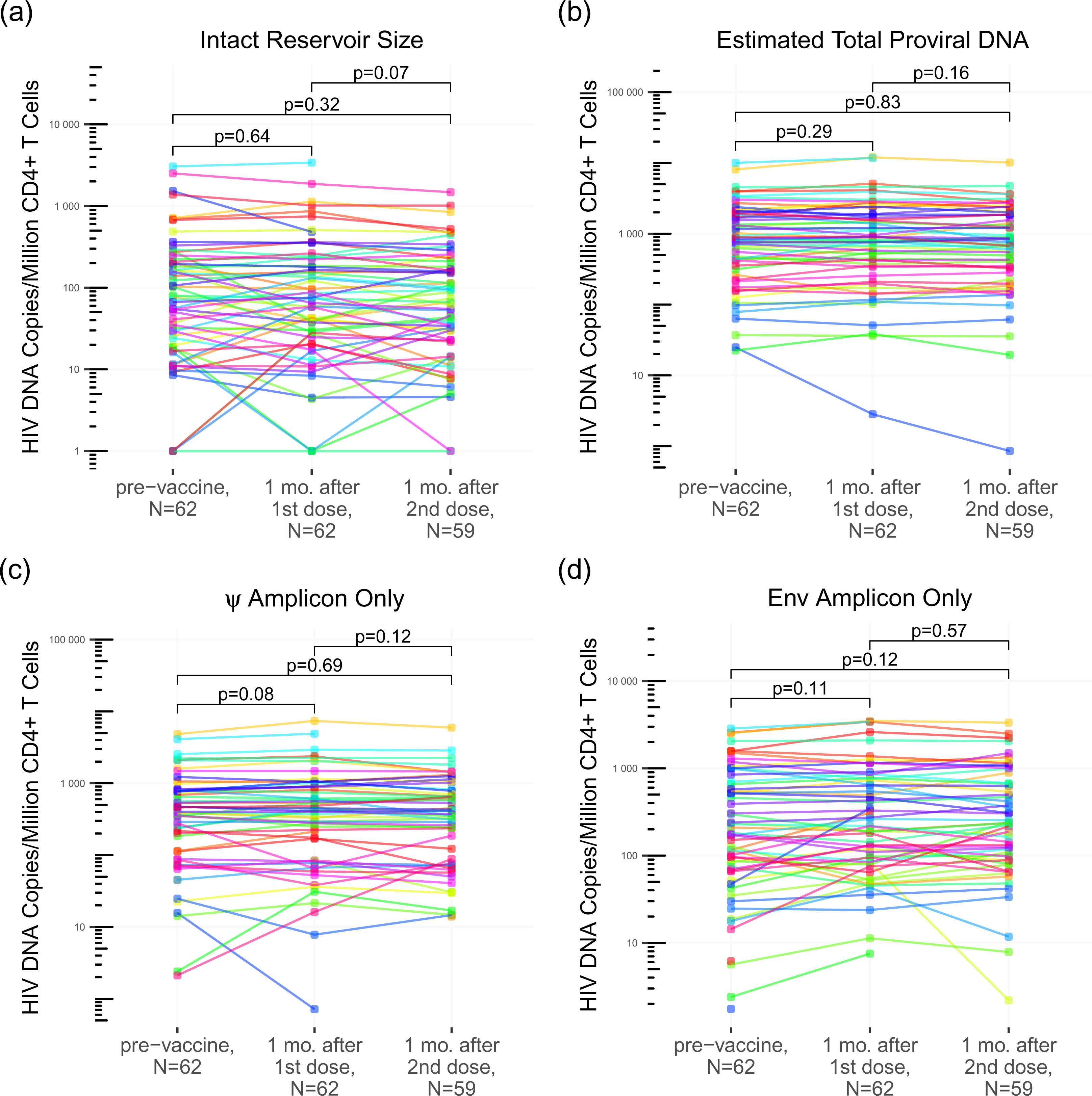
Measures of intact reservoir size, and total, 5’-defective, 3’-defective proviral burdens after one- and two-dose COVID-19 vaccination. Intact reservoir size (*panel a*), total proviral burden (*panel b*), 5’-defective proviral burden (panel *c*), and 3’-defective proviral burden (*panel d*) measured at baseline (pre-vaccine), one month after the first vaccine dose, and one month after the second vaccine dose, using the Intact Proviral DNA Assay (IPDA). Each participant is identified by a unique color that is consistent throughout all figures. P-values were calculated using the Wilcoxon sum rank test for paired data.

### No evidence that the magnitude of the COVID-19-vaccine-induced immune response increases the likelihood of HIV reservoir perturbation

Based on the observation that PWH on ART who mounted strong responses to influenza vaccination were more likely to show transient increases in HIV pVL [20], we investigated whether the magnitude of the COVID-19-vaccine-induced immune response increased the likelihood of HIV reservoir perturbation. We found no evidence to support this: one month following the first vaccine dose, anti-SARS-CoV-2-Spike serum antibody concentrations were a median of 51.4 (19.4-130.6 U/mL) in participants who maintained pVL <20 copies/mL versus a median of 36.6 (16.4-80.7 U/mL) among participants with a pVL >20 HIV copies/mL, a difference that was not statistically significant (Mann-Whitney p = 0.73; **Fig. 3a**). Similarly, one month after the second vaccine dose, anti-SARS-CoV-2-Spike serum antibody concentrations were median of 8970 (5019-13544 U/mL) and 7205 (4163-11638 U/mL) respectively in participants with pVL <20 versus >20 copies/mL, a difference that was not statistically significant (p=0.84; **Fig. 3b**).

**Figure 3.**
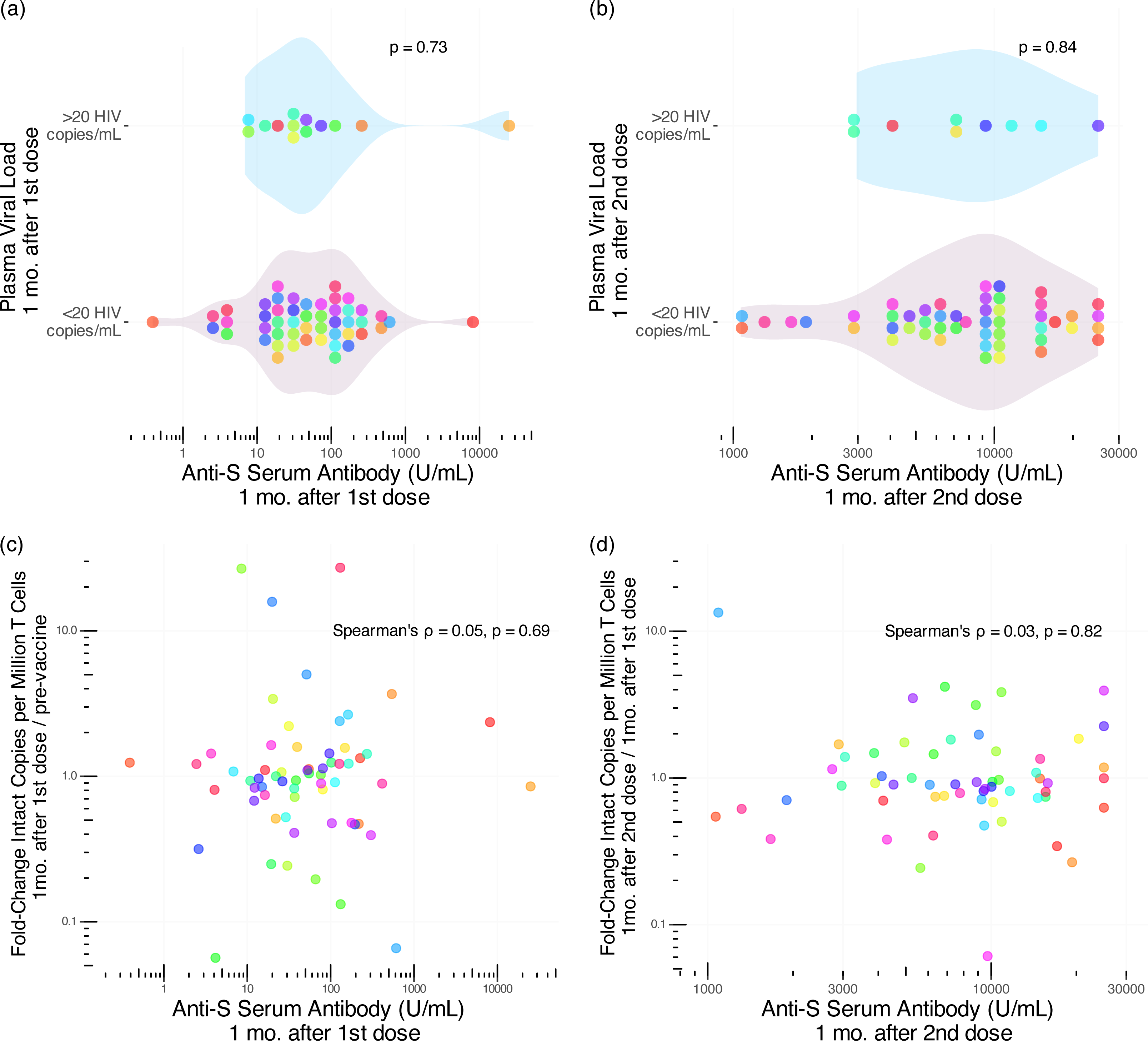
Relationship between reservoir size, plasma viral load and COVID-19 vaccine immune response magnitude. *Panel a:* Anti-SARS-CoV-2-Spike (S) serum antibody levels one month following the first COVID-19 vaccine dose in participants with pVL >20 HIV RNA copies/mL (top group) versus <20 HIV RNA copies/mL (bottom group) at this time point. P-value calculated using the Mann-Whitney U-test. *Panel b*: same as *a*, but for anti-S serum antibody levels and pVL one month after the second dose. *Panel c:* Relationship between Anti-S serum antibody levels one month following the first COVID-19 vaccine dose, and the fold-change in intact reservoir size from baseline, assessed using Spearman’s correlation. Panel *d*: same as *c*, but depicting the relationship between Anti-S serum antibody levels one month following the second COVID-19 vaccine dose, and the fold-change in intact reservoir size since the previous time point. Each participant is identified by a unique color that is consistent throughout all figures.

Likewise, the magnitude of anti-SARS-CoV-2-Spike serum antibody levels one month after the first vaccine dose did not correlate with the fold-change in reservoir size from baseline (Spearman ρ=0.05, p=0.69; Fig. 3c), nor did the magnitude of anti-SARS-CoV-2-Spike serum antibody levels one month after the second vaccine dose correlate with the fold-change in reservoir size from the prior visit (Spearman ρ=0.03, p=0.82; Fig. 3d). Both the pVL and reservoir size results remained consistent after excluding the participants with prior COVID-19 (all p>0.51; not shown). Overall, the results from our observational cohort do not provide any evidence that COVID-19 mRNA vaccines modulated the HIV reservoir nor induced plasma viremia.

### Province-wide analysis of trends in HIV plasma viral loads before and after mass COVID-19 vaccination

We next investigated whether BC’s mass COVID-19 vaccination campaign was associated with an increase in the frequency of detectable pVL test results (defined here as pVL >40 copies HIV RNA/mL; see methods) at the population level. We began by analyzing all 290,401 pVL tests performed in BC since 2012. These represented all pVL tests performed as part of routine clinical care of all PWH in BC, regardless of the individual’s ART status at the time of testing (the number of PWH undergoing pVL testing in a given year ranged from 7112-8095 during this period) (**Fig. 4a**). Between 2012 and approximately 2016, the percentage of detectable pVL test results declined from nearly 29% to an average of 16% as a result of a province-wide implementation of widespread HIV testing and immediate initiation of free-of-charge ART that began in 2013 [41]. Since 2016, the overall percentage of detectable pVL has remained relatively stable, though there was a slight uptick in the proportion of detectable pVL tests during 2020 because care providers were asked to reduce the frequency of routine pVL testing for PWH with long-term viremia suppression, to preserve lab capacity for COVID-19 diagnostic testing (which was also performed on the cobas 6800 in BC).

**Figure 4.**
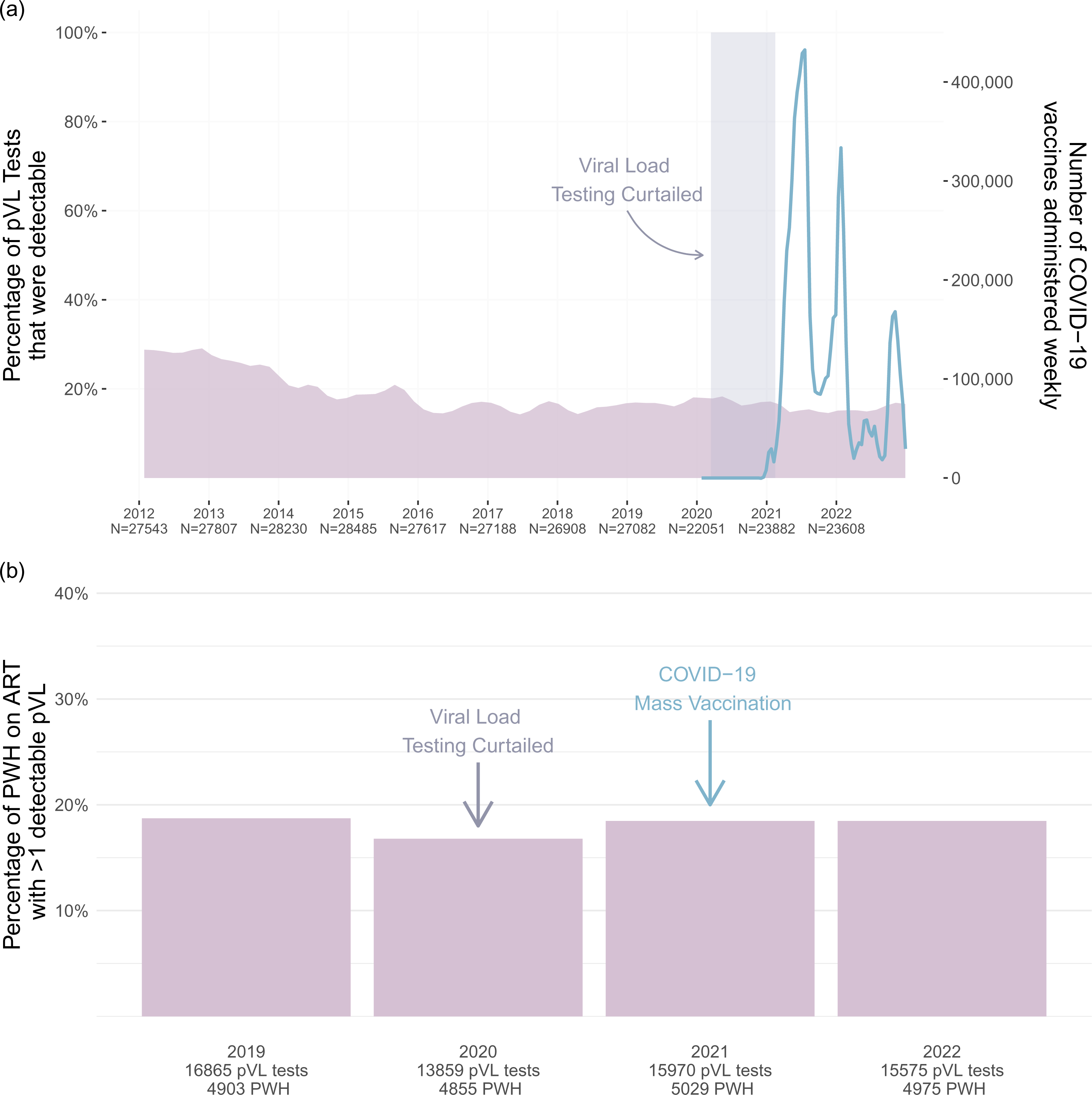
Population-level analysis of pVL test results in BC. *Panel a*: The percentage of all pVL tests performed in BC that returned a detectable result (defined as >40 HIV RNA copies/mL; see methods) was computed for every month between 2012-2022 and depicted as a smoothed mauve curve. The numbers underneath the x-axis denote the total number of pVL tests performed each year. The smoothed blue line depicts the number of COVID-19 vaccine doses administered in BC weekly, as retrieved from the COVID-19 Vaccine Tracker [36]. The shaded rectangle denotes the period when pVL load testing of long-term ART-suppressed PWH was temporarily curtailed to preserve capacity for COVID-19 diagnostic testing. *Panel b*: The percentage of PWH in BC receiving uninterrupted ART who experienced at least one detectable pVL measurement (defined as >40 HIV RNA copies/mL; see methods) for the given year. The total number of pVL tests included in each year’s analysis, as well as the total number of PWH from whom these pVL data are derived, are shown on the x-axis.

COVID-19 vaccines were first made available to priority populations in BC, namely frontline health workers, long-term care residents and individuals with select clinical conditions (which did <u>not</u> include HIV infection) starting in late December 2020. The age-based mass immunization campaign began in April 2021, which is when the majority of PWH became eligible for COVID-19 vaccination. Peak administration of first doses occurred from approximately May through September 2021, with second doses largely administered from February to March 2022 (**Fig. 4a**). By April 10^th^, 2022, 89% of all adult British Columbians had received at least two COVID-19 vaccine doses [35]. Notably, we observed no obvious evidence of population-level increases in detectable pVL during or immediately following the peak vaccine administration periods in the province (**Fig. 4a**).

As transient, vaccine-induced viremia may only be observable in PWH on suppressive ART, we next restricted the province-wide analysis to PWH receiving uninterrupted ART, defined as those with continuous monthly prescription refill records in a given year. We then determined the annual percentage of PWH receiving uninterrupted ART who experienced at least one detectable pVL that year. We began the analysis in 2019, the year before the COVID-19 pandemic was declared (and the first full year that the cobas 6800 HIV Test was implemented) (**Fig. 4b**). Between 4903 and 5975 PWH were included in each year’s analysis. In 2019, 18.7% of PWH receiving uninterrupted ART experienced at least one detectable pVL. In 2020, the percentage was 16.8%, though this reduction may be attributable to the temporary reduction in pVL testing that year. By 2021, pVL testing returned to pre-pandemic levels and >9 million COVID-19 vaccine doses were administered provincewide in that year. Nevertheless, the percentage of PWH receiving uninterrupted ART who experienced at least one detectable pVL measurement was 18.5%, which was essentially identical to that observed in 2019 prior to the pandemic. The percentage for 2022 was also 18.5%. Taken together, these results reveal no evidence linking the provincial COVID-19 vaccination campaign to increases in detectable pVL at the population level in BC.

## Discussion

The mass rollout of COVID-19 mRNA vaccines provided an opportunity to study the potential stimulatory effects of this new vaccine modality on the HIV reservoir. Though a number of studies have now investigated this topic [27–29,31], their results have not been entirely conclusive. While one study found evidence of a gradual, though not statistically significant increase in the rate of detectable viremia peaking 4 weeks after the second vaccine dose [27], two others reported no changes in viremia following two-dose vaccination [28,30]. Two studies reported reduced frequencies of detectable viremia after three-dose vaccination [28,29], though in one study this was likely attributable to increased time on ART, which was initiated around the study outset for many participants [28]. Ours is the first study to our knowledge to combine cohort- and population-level analyses of pVL trends during a mass COVID-19 mRNA vaccination campaign.

Overall, neither the cohort nor population-level analyses identified evidence that COVID-19 mRNA vaccination promotes viral release from the reservoir to detectable levels in plasma. One month after the first and second vaccine doses, the proportion of participants with detectable pVL remained statistically unchanged from baseline, with no participant experiencing virologic failure (defined as pVL>200 copies/mL [39,40]).

Moreover, there was no evidence that the magnitude of the anti-SARS-CoV-2-Spike antibody response influenced the likelihood of experiencing plasma viremia post-vaccination. Our province-wide analysis similarly found that the frequency of detectable pVL test results remained stable at the population level following the mass administration of first, second and booster COVID-19 vaccine doses in BC. This remained the case whether we considered the overall PWH population, or the subset receiving uninterrupted ART.

Our cohort-based analysis also found no evidence that COVID-19 mRNA vaccination induced changes in HIV reservoir size, nor in total, 5’-defective, or 3’-defective proviral loads. This is consistent with findings from of three studies that assessed smaller numbers of participants for this outcome using similar approaches [27,29,31].

Our study has some limitations and caveats. We sampled our cohort one month following each COVID-19 vaccine dose because our primary objective was to evaluate vaccine immune responses in PWH, as previously reported [6]. This timing however would have missed rapid viremia events that had resolved by this time (indeed, such rapid viremia events have been reported for influenza vaccination [20]). That said, in reports describing viremia following COVID-19 vaccination, including the single case study, viremia was detectable one month post-vaccination [25,27]. Another limitation is that our province-wide evaluation represents an ecological analysis that correlated population-level pVL and vaccination data, because COVID-19 vaccination dates of individual British Columbians were not available to us. Thus, even though this analysis captured all HIV pVL tests performed in BC during the period of interest, the variable timing of these tests with respect to the individual’s vaccination date would not have allowed us to capture all viremia events that may have occurred. Finally, though neither our cohort nor population-level analyses support frequent nor widespread effects of COVID-19 vaccination on the HIV reservoir, we cannot rule out that such events may occur uncommonly, through mechanisms that remain incompletely understood.

In conclusion, we found no evidence that COVID-19 mRNA vaccines induced changes in HIV reservoir size nor plasma viremia in PWH receiving suppressive ART. Taken together with similar findings from other studies [27–29,31], we conclude that there is now a strong body of evidence indicating COVID-19 immunization is safe and effective in PWH [4–12]. This should provide additional reassurance to PWH and their care providers regarding the safety of COVID-19 mRNA vaccines.

## Data Availability

All data produced in the present study are available upon reasonable request to the authors

## Acknowledgements

MCD and ZLB conceived and designed the study. MAB and ZLB obtained funding. HRL, MH, MAB and ZLB established the cohort. MCD, FHO, NNK, SS and NM-G processed specimens and/or collected IPDA data. MCD analyzed data and made figures. TL, CFL and MGR performed the plasma viral load testing. MLD, JS, DTH and MGR performed the COVID-19 serologic testing. NB, PS and CJB performed the province-wide viral load analysis. JSGM and RB provided data access. MCD wrote the first draft of the paper, with all authors contributing edits.

We are grateful to the study participants, without whom this research would not be possible.

This work was supported in part by the Canadian Institutes for Health Research (CIHR) through a project grant (PJT-159625 to ZLB) and a team grant (HB1-164063 to ZLB and MAB). This work was also supported in part by the National Institutes of Health (NIH) through the Martin Delaney “REACH” Collaboratory (NIH grant 1-UM1AI164565-01 to ZLB and MAB), which is supported by the following NIH cofounding institutes: NIMH, NIDA, NINDS, NIDDK, NHLBI and NIAID. MCD was supported by a CIHR CGS-M award. NNK was supported by a CIHR Vanier Canada Graduate Scholarship. MLD and ZLB were supported by Scholar Awards from Michael Smith Health Research BC. The content is solely the responsibility of the authors and does not necessarily represent the official views of the National Institutes of Health or other funders.

